# Interrelations between Atherogenic Index of Plasma, Subclinical Myocardial Injury, and Cardiovascular Mortality in the General Population

**DOI:** 10.64898/2026.01.14.26344126

**Authors:** Uttsav B. Sandesara, Richard Kazibwe, Joseph Yeboah, Elsayed Z. Soliman

## Abstract

**Objective:** To examine the association between Atherogenic Index of Plasma (AIP) and subclinical myocardial injury (SCMI), and their combined impact on cardiovascular disease (CVD) mortality in the general population.

**Methods:** This analysis included 7,093 participants without CVD from the Third National Health and Nutrition Examination Survey. AIP was calculated as the logarithmic ratio of triglycerides to HDL cholesterol. Participants were stratified into low or high AIP groups based on the median AIP value (0.958). Electrocardiographic SCMI was defined as Cardiac Infarction/Injury Score ≥10 points. CVD mortality data were obtained from the National Death Index. Multivariable logistic regression models assessed the baseline cross-sectional association between AIP and SCMI, while Cox proportional hazards models examined the relationship between different baseline AIP/SCMI groups and CVD mortality.

**Results:** High AIP was associated with increased odds of SCMI [OR(95% CI): 1.20(1.07–1.35)] in multivariable logistic regression analysis. In multivariable Cox proportional hazard models, compared to participants with low AIP and absent SCMI, those with SCMI had a higher risk of CVD mortality regardless of AIP level [(HR(95% CI) 1.28(1.05–1.57) and 1.33(1.10–1.60)]. However, high AIP without SCMI was not associated with CVD mortality [HR (95% CI) 0.94(0.79–1.11)].

**Conclusions:** High AIP was associated with an increased risk of SCMI. SCMI was linked to a higher risk of CVD mortality regardless of AIP levels, while high AIP was only associated with CVD mortality when SCMI was present, suggesting that the reported adverse outcomes linked to high AIP may be driven by the development of SCMI.

## Introduction

Cardiovascular disease (CVD) remains the leading cause of morbidity and mortality worldwide, representing a significant global health burden [1, 2]. Early identification of individuals at high risk of CVD before clinical manifestation is essential for reducing adverse outcomes [3]. The Atherogenic Index of Plasma (AIP)—calculated as the logarithmic ratio of triglycerides to high-density lipoprotein cholesterol (HDL-C)—has emerged as a novel biomarker linked to adverse cardiovascular outcomes [4, 5]. Although traditional lipid measures, such as non-HDL-C, effectively capture the total burden of atherogenic lipoproteins, AIP provides additional biological insights. Specifically, AIP has the advantage of addressing the intrinsic variability and skewed distribution characteristic of triglyceride values, enhancing measurement stability. Biologically, AIP uniquely correlates with concentrations of small, dense LDL (sdLDL) particles, which have demonstrated greater atherogenic potential compared to larger LDL particles [6–8]. Thus, AIP offers a dynamic measure of lipid metabolism by capturing the key balance between pro-atherogenic triglyceride-rich lipoproteins and protective HDL particles, potentially serving as an early and more informative predictor of cardiovascular events than traditional lipid parameters alone [8, 9].

Subclinical myocardial injury (SCMI), detected using validated electrocardiographic tools such as the Cardiac Infarction/Injury Score (CIIS), represents another emerging fundamental marker in cardiovascular risk assessment [10]. SCMI, defined as CIIS ≥10 score points, has shown to reflect early myocardial stress and damage, often in the absence of overt clinical symptoms, and is associated with an increased risk of cardiovascular mortality in individuals without diagnosed CVD [11].

Currently, the relationship between AIP and SCMI remains unclear, with no previous literature explicitly exploring their potential interactions or combined influence on cardiovascular outcomes. Given the unique biological insights provided by AIP into lipid metabolism and the sensitivity of SCMI for detecting early myocardial damage, clarifying their relationship could enhance early identification of individuals at heightened cardiovascular risk. Understanding whether AIP and SCMI are interrelated, as well as whether their combined presence confers an elevated risk for cardiovascular mortality, is critical for improving risk stratification and preventive strategies. Therefore, we aimed to explore the potential interplay between AIP and SCMI and to assess their combined impact on cardiovascular mortality.

## Methods

### Study Design and Population

We utilized publicly accessible data from the Third National Health and Nutrition Examination Survey (NHANES-III), a survey of nationally representative cohort of non-institutionalized individuals in the United States conducted between 1988 and 1994 [12]. The survey was approved by the National Center for Health Statistics (NCHS) Research Ethics Review Board and informed consent was provided by all participants. Previous studies have outlined the design and components of NHANES-III [13].

For this study, we only included participants from NHANES-III who underwent an ECG recording (n=8561) and had complete data to assess AIP (triglycerides and HDL-C). We excluded participants with a history of clinical CVD, incomplete ECG data, missing triglyceride and HDL-C levels, and missing baseline characteristic data. Following exclusions, a total of 7,093 individuals were included in the final analysis.

### Atherogenic Index of Plasma (AIP)

Blood samples, collected via venipuncture, were obtained in the fasting state for analysis of total HDL-C and TG among other components as reported by the NCHS [14]. AIP was calculated as the logarithmic ratio of triglycerides to HDL-C: log10(triglycerides [mmol/L]/HDL-C [mmol/L]) [6]. We evaluated AIP both as a continuous variable and a categorical variable to assess its association with SCMI and CVD mortality.

We utilized AIP as a continuous variable as demonstrated in prior studies [4–6]. For categorical analysis, participants were stratified into two groups based on the median AIP value (0.958) within the cohort, as modeled in prior studies [7–9]. Those with AIP values above the median (>0.958) were classified as having high AIP, while those below the median (≤0.958) were classified as having low AIP.

### Subclinical Myocardial Injury (SCMI)

SCMI was defined using the CIIS, a validated electrocardiographic measure based on a weighted scoring system that incorporates several electrocardiographic waveform components. Specifically, the CIIS in the NHANES-III database was calculated using the multivariate electrocardiographic scoring system derived from a combination of 11 discrete ECG features (binary or ternary thresholds) and 4 continuous ECG measurements obtained from standard 12-lead ECGs (including two inverted leads, -aVR and -aVL) [10, 11]. Each ECG feature was assigned empirically derived weights, optimized through multivariate discriminant analysis to detect myocardial injury [10, 11]. In NHANES-III, CIIS was computed via automated computerized analysis at a central ECG core laboratory, with subsequent visual inspection of outlier values by trained technicians [11]. CIIS values provided in NHANES-III were originally multiplied by 10 to avoid decimal points and were divided by 10 prior to analysis, reflecting standardized, validated measurements rather than solely manual human scoring [11]. Consistent with previous studies, SCMI was defined as a CIIS score of ≥10, reflecting significant myocardial stress and damage that might not be detectable by traditional clinical methods[10, 11].

### Group Stratification of AIP and SCMI

To assess the combined impact of AIP and SCMI with CVD mortality, participants were categorized into four distinct groups based on their AIP (high versus low, using the median as the cutoff) and SCMI (presence or absence) status: Low AIP/absent SCMI (reference group), low AIP/present SCMI, high AIP/absent SCMI, and high AIP/present SCMI.

### Cardiovascular Mortality

CVD mortality was ascertained by linking NHANES III participants to the National Death Index (NDI) through a detailed matching process using multiple personal identifiers, such as social security number, sex, and date of birth. CVD deaths were classified according to ICD-10 codes 100–178. Mortality data for the NHANES III cohort were available up until December 31, 2019.

### Covariates

Sociodemographic information, including age, sex, race/ethnicity, education level and smoking status, was collected via self-report during an in-home interview. Obesity was defined as a body mass index (BMI) of 30 kg/m² or higher. Hypertension was identified as either a systolic blood pressure ≥130 mmHg, diastolic blood pressure ≥80 mmHg, or the use of antihypertensive medications. Diabetes was defined as a fasting blood glucose level of 126 mg/dL or higher. Physical activity was assessed based on participants’ self-reported frequency, type, and intensity of leisure-time activities.

### Statistical Analysis

Multivariable logistic regression models were used to evaluate the cross-sectional associations between Atherogenic Index of Plasma (AIP) and subclinical myocardial injury (SCMI). AIP was analyzed both as a categorical variable (high vs. low, based on the median) and as a continuous variable (per standard deviation [SD] increase). Odds ratios (ORs) with 95% confidence intervals (CIs) were calculated to evaluate these associations. Model 1 was adjusted for age, sex, race/ethnicity, and education level, while Model 2 included additional adjustments for clinical covariates such as diabetes, systolic blood pressure, antihypertensive medication use, body mass index (BMI), use of lipid-lowering medications, smoking status, and physical activity. These covariates were selected based on their known associations with cardiovascular outcomes and lipid metabolism.

Cox proportional hazards models were used to examine the relationship between the combined AIP/SCMI groups and CVD mortality. Hazard ratios (HRs) with 95% CIs were calculated for the four AIP/SCMI groups: Low AIP/absent SCMI, low AIP/present SCMI, high AIP/absent SCMI and high AIP/present SCMI. The group with low AIP and absent SCMI served as the reference group. Model 1 was adjusted for age, sex, race/ethnicity, and education level, while Model 2 included further adjustments for diabetes, systolic blood pressure, antihypertensive medications, BMI, lipid-lowering medications, smoking, and physical activity.

To assess whether the association between AIP and CVD mortality was modified by the presence of SCMI, interaction terms between AIP and SCMI were tested. We performed these interaction analyses using three approaches: (1) treating both AIP and SCMI as continuous variables, (2) treating both AIP and SCMI as categorical variables, and (3) analyzing AIP as a continuous variable while SCMI was treated as categorical. Each interaction model was adjusted for the same covariates included in Model 2.

For comparative purposes, we also evaluated non-HDL-C levels, calculated as total cholesterol minus HDL-C, to assess its association with SCMI and CVD mortality. Non-HDL-C was analyzed similarly to AIP, both categorically (high vs. low, based on the cohort median) and continuously (per 1mg/dL and SD increase). These supplemental analyses are presented in detail in the Supplemental Material.

All statistical analyses were performed using SAS version 9.4 (SAS Institute Inc., Cary, NC). Two-sided p-values of less than 0.05 were considered statistically significant.

## Results

### Baseline Characteristics

A total of 7,093 participants free of CVD were included in the analysis (age 59.3± 13.4 years, 52.8% women, 49.4% White). **Table 1** show the baseline characteristics of the study participants stratified by AIP and SCMI status. A total of 1,861 participants (26.3%) had SCMI, and 3,533 had high AIP. Participants with both SCMI and AIP tended to be older and had more prevalent CVD risk factors than those without those two conditions.

**Table 1:**
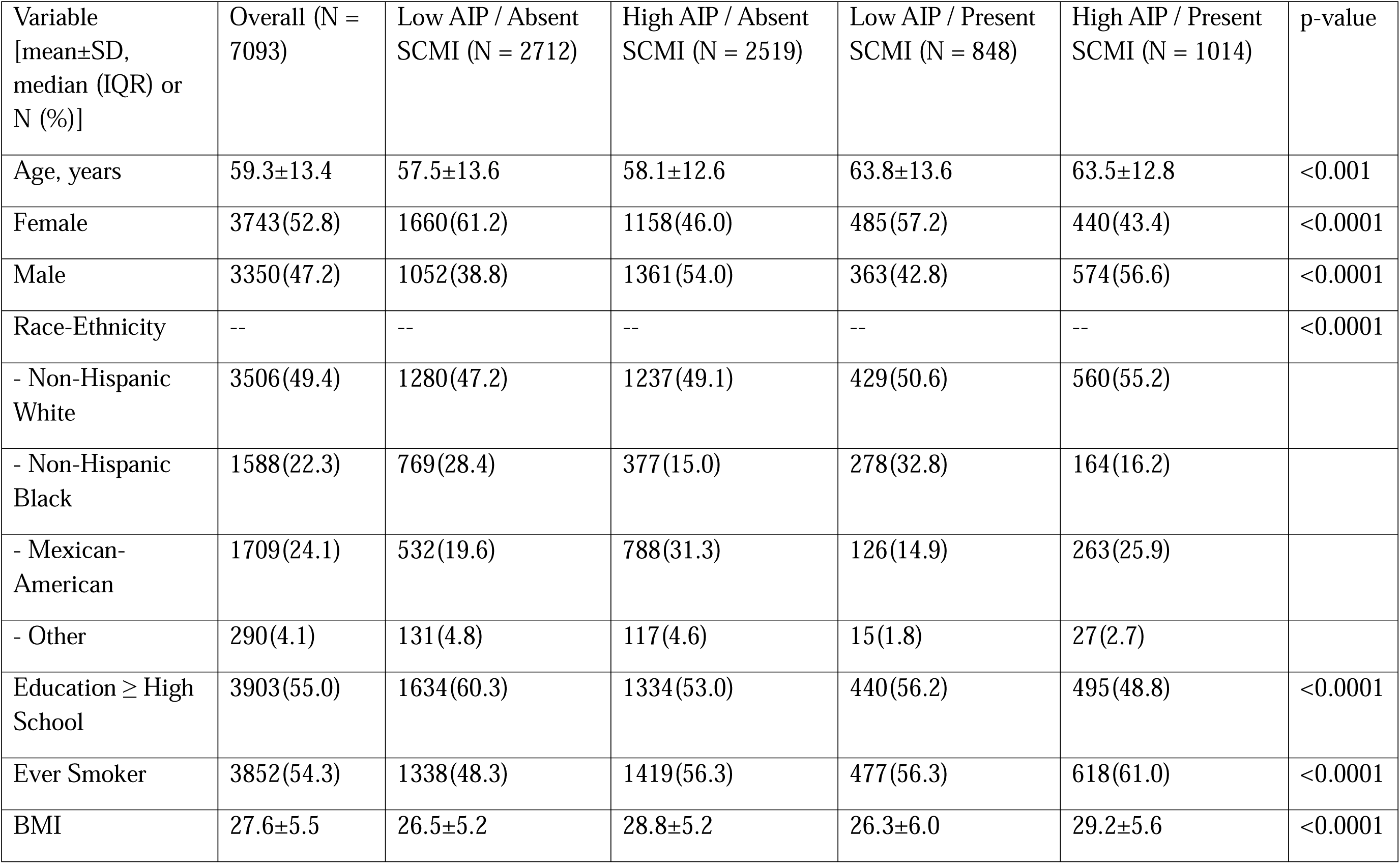
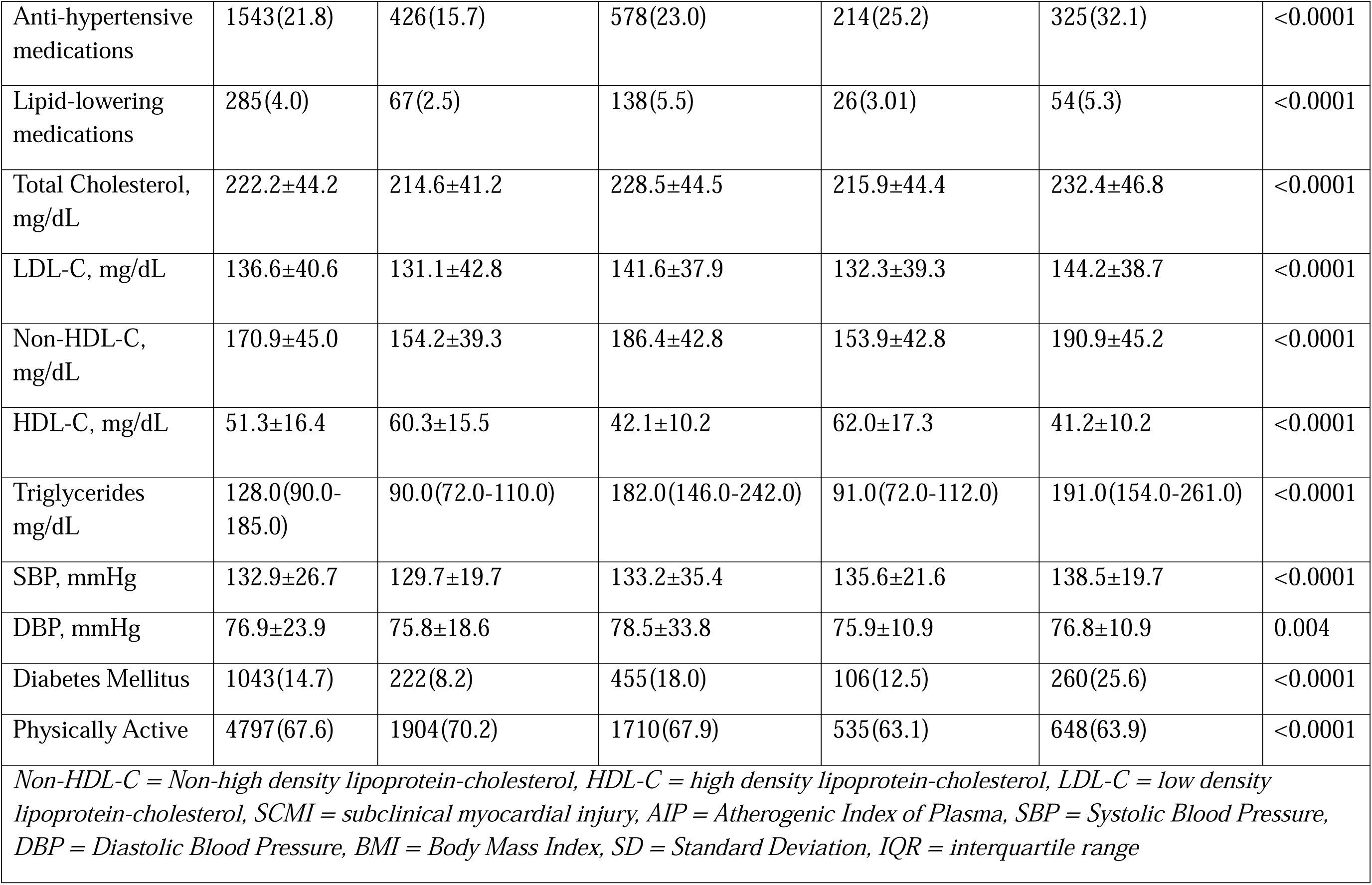
Baseline Characteristics by AIP and SCMI Status.

### Association of AIP with SCMI

In the cross-sectional analysis, high AIP was significantly associated with an increased likelihood of having SCMI. In a model adjusted for socio-demographics, participants with high AIP had 26% (p<0.001) greater odds of having SCMI compared to those with low AIP. This association remained significant after adjusting for additional CVD risk factors, with participants having high AIP demonstrating 20% (p=0.002) higher odds of SCMI. In a similar model, each standard deviation (SD) increase in AIP was associated with 19% (p<0.001) greater odds of SCMI in fully adjusted models (**Table 2)**.

**Table 2:** Baseline Association of AIP with SCMI.

|  | Model 1 |  | Model 2 |  |
| --- | --- | --- | --- | --- |
|  | OR (95%CI) | *P-value* | OR (95%CI) | *P-value* |
| **Categorical** |  |  |  |  |
| Low AIP | Reference | -- | Reference | -- |
| High AIP | 1.26 (1.13–1.40) | <0.0001 | 1.20 (1.07–1.35) | 0.002 |
| **Continuous** |  |  |  |  |
| AIP (continuous) | 1.26 (1.18 – 1.37) | <0.0001 | 1.19 (1.09 – 1.28) | <0.0001 |
| AIP (per SD) | 1.20 (1.13 – 1.27) | <0.0001 | 1.14 (1.07 – 1.21) | <0.0001 |
| *OR, Odds Ratio; CI, confidence interval*<br>**Model 1** adjusted for age, sex, race and education level.<br>**Model 2** adjusted for model 1 plus diabetes, systolic BP, antihypertension medication, body mass index, lipid lowering medications, smoking, and physical activity. |  |  |  |  |

### Association of AIP and SCMI with CVD Mortality

In a Cox proportional hazards model adjusted for socio-demographics, participants with both high AIP and SCMI had a 56% (p<0.001) higher risk of CVD mortality compared to the reference group (low AIP and absent SCMI). This association remained significant after further adjustment for CVD risk factors, with a 33% (p = 0.004) higher risk of CVD mortality. In similar models, participants with SCMI but low AIP also had a 40% (p<0.001) and 28% (p = 0.014) elevated risk of CVD mortality compared to the reference group. On the other hand, the associations of high AIP without SCMI with CVD mortality did not reach statistical significance in either of the models (**Table 3**). There was no significant interaction between AIP and SCMI with respect to CVD mortality (interaction p-value = 0.477).

**Table 3:** Association of AIP/SCMI Status with CVD mortality.

| AIP and SCMI Status | Participants (n)/ Events (%) | Model 1 |  | Model 2 |  |
| --- | --- | --- | --- | --- | --- |
|  |  | HR (95%CI) | P-value | HR (95%CI) | P-value |
| SCMI Absent + Low AIP | 2,712/10.6 | Reference | -- | Reference | -- |
| SCMI Absent + High AIP | 2,519/10.8 | 0.71 (1.03 – 1.22) | 0.709 | 0.94 (0.79 – 1.11) | 0.447 |
| SCMI Present + Low AIP | 848/17.9 | 1.40 (1.15 – 1.71) | <0.001 | 1.28 (1.05 – 1.57) | 0.014 |
| SCMI Present + High AIP | 1,014/20.4 | 1.56 (1.32 – 1.90) | <0.0001 | 1.33 (1.10 – 1.60) | 0.004 |
| *SCMI = subclinical myocardial injury, AIP = Atherogenic Index of Plasma, CVD = cardiovascular disease, HR = Hazard Ratio, CI = Confidence Interval*<br>**Model 1** adjusted for age, sex, race and education level.<br>**Model 2** adjusted for model 1 plus history of diabetes, systolic blood pressure, use of antihypertension medication, body mass index, use of lipid lowering medications, smoking status, and physical activity. |  |  |  |  |  |

### Exploratory Analysis of Association of Non-HDL-C, SCMI and CVD Mortality

In supplemental exploratory analyses, elevated non-HDL-C was not independently associated with higher odds of SCMI after adjusting for socio-demographic and CVD risk factors (OR 0.96 (0.83-1.12), p = 0.606). Participants with both SCMI and elevated non-HDL-C had a 29% (p = 0.044) higher risk of cardiovascular mortality compared to those without these conditions, after adjusting for socio-demographic and CVD risk factors **(Supplemental Tables 1–2)**.

## Discussion

In this analysis from NHANES III, we explored the interrelationship between the Atherogenic Index of Plasma (AIP), subclinical myocardial injury (SCMI), and CVD mortality in a population without clinically diagnosed CVD. There are three key findings from our study: 1) Individuals with high levels of AIP are at an increased risk of SCMI; 2) Individuals with both SCMI and high levels of AIP are at increased risk of CVD mortality than those without these conditions; 3) Irrespective of AIP level, SCMI is associated with a higher risk of CVD mortality. Overall, these findings suggest that the adverse outcomes linked to high AIP may be driven by the development of SCMI.

AIP has emerged as a valuable biomarker for assessing lipid abnormalities and their contributions to atherogenesis, reflecting an imbalance between pro-atherogenic lipids and protective HDL cholesterol [10, 11]. Our use of AIP over guideline-recommended non-HDL cholesterol was intentional, grounded in its unique pathophysiologic relevance. While non-HDL-C broadly captures all ApoB-containing lipoproteins, prior evidence supports AIP’s enhanced predictive value for certain cardiovascular outcomes, particularly due to its close association with the more atherogenic sdLDL particle leading to insulin resistance, metabolic syndrome, and lipid metabolism derangements that are not fully represented by non-HDL cholesterol alone [5–8, 15–17]. Our findings align with these studies, demonstrating AIP’s potential as an early marker for myocardial injury and cardiovascular events. Although high AIP did not independently predict CVD mortality, it was associated with an increased risk of SCMI, highlighting AIP’s role in detecting early myocardial injury and subclinical cardiovascular risk. We have included additional exploratory analyses (see Supplemental Material) assessing the association of non-HDL-C, SCMI and CVD mortality. While non-HDL-C alone was not associated with SCMI or CVD mortality, its combination with SCMI identified a subgroup at significantly higher risk, suggesting that structural and lipid-based markers may provide complementary risk stratification (Supplemental Tables 1–2).

SCMI, detected using the CIIS, has been validated as a marker for subclinical myocardial damage[10, 11, 14, 18–21]. Our findings confirm that SCMI was independently associated with higher cardiovascular mortality, underscoring the critical role of early myocardial injury in driving adverse outcomes. The use of CIIS demonstrates its value in detecting myocardial damage before clinical symptoms appear, making it an effective tool for early risk stratification.

Although the interaction between AIP and SCMI was not statistically significant, the presence of both risk factors was associated with a higher risk of CVD mortality. A plausible biological causal chain is that elevated AIP, reflecting atherogenic lipid imbalance, initiates atherosclerosis and subsequent myocardial injury through mechanisms involving inflammation, oxidative stress, and endothelial dysfunction. This process might explain the observed collinearity and attenuation of AIP’s significance in models that simultaneously include SCMI. Therefore, while elevated AIP may represent an initial contributor to myocardial injury, the presence of SCMI could mediate AIP’s direct association with cardiovascular mortality. This suggests that AIP and SCMI operate through complementary pathways, each contributing to the overall risk. The combination of these two markers may identify individuals at particularly high risk for cardiovascular events, but further longitudinal research is needed to further clarify these relationships and explore their long-term impact.

The combined use of AIP and SCMI offers a practical approach to early risk stratification in clinical settings. AIP is an accessible and cost-effective biomarker for lipid imbalances, while SCMI, assessed through ECG-based CIIS, is a non-invasive method for identifying subclinical myocardial damage. Incorporating these markers into routine care could help identify high-risk individuals, particularly those with dyslipidemia or early myocardial injury, and support personalized interventions such as lipid-lowering therapies or lifestyle modifications [22–24]. The advantage of CIIS is that it is easily obtained from a standard 12-lead ECG, making it a cost-effective diagnostic tool. For underserved populations with limited access to cardiovascular diagnostics, using simple tools like ECG and CIIS for routine screening could improve early detection and intervention. Future studies may explore the potential utility of artificial intelligence (AI)-enhanced ECG interpretation in further refining and standardizing the detection of SCMI.

Our study’s strengths include the use of a large, nationally representative sample from NHANES III, which provided a diverse population with long-term follow-up data on cardiovascular outcomes. This robust sample allowed for a detailed analysis of the relationships between AIP, SCMI, and CVD mortality, making the findings potentially generalizable. However, our study also has some limitations. First, despite adjusting for numerous potential confounders, residual confounding cannot be entirely ruled out as some participants may have had undiagnosed CVD at baseline. Second, AIP and SCMI were measured at a single time point, limiting our ability to assess how changes in these markers affect risk over time. Finally, this is a cross-sectional analysis, and while it provides valuable associations, it does not allow for the determination of causality between AIP, SCMI, and cardiovascular mortality.

## Conclusions

In conclusion, our study demonstrates that high AIP is associated with increased risk of SCMI, and that participants with both conditions are at a higher risk of CVD mortality than those without these conditions. Irrespective of AIP level, SCMI was associated with a higher risk of CVD mortality suggesting that the adverse outcomes linked to high AIP may be driven by the development of SCMI. Incorporating both AIP and SCMI into cardiovascular risk assessments could enhance early detection and guide targeted interventions, particularly in individuals without clinically apparent disease. Future studies are essential to confirm these associations and explore whether early interventions based on these markers can reduce cardiovascular mortality.

## Data Availability

The datasets analyzed for this study are publicly available from the National Health and Nutrition Examination Survey (NHANES): https://wwwn.cdc.gov/nchs/nhanes/nhanes3/default.aspx. The present study involved a secondary analysis of these data; no new human subject data were generated.Derived variables and analytic code are available from the corresponding author upon reasonable request.

## Data Statement

The data supporting this study are openly available in NHANES-III at https://wwwn.cdc.gov/nchs/nhanes/default.aspx.

## Ethics Approval

NHANES III was approved by the National Center for Health Statistics Research Ethics Review Board.

## Patient Consent

Documented consent was obtained from all participants as part of NHANES-III.

## CRediT authorship contribution statement

Uttsav Sandesara: Writing – original draft, visualization. Richard Kazibwe: Data curation, formal analysis, visualization. Joseph Yeboah: Writing – review and editing. Elsayed Soliman: Writing – review and editing, supervision

## Supplemental Material

**Supplemental Table 1:** Association of Non-HDL Cholesterol with Subclinical Myocardial Injury.

|  | **Model 1** |  | **Model 2** |  |
| --- | --- | --- | --- | --- |
|  | **OR (95%CI)** | ***P-value*** | **OR (95%CI)** | ***P-value*** |
| **Categorical** |  |  |  |  |
| Low non-HDL-C | Reference | -- | Reference | -- |
| High non-HDL-C | 0.99 (0.86-1.15) | 0.935 | 0.96 (0.83-1.12) | 0.606 |
| **Continuous** |  |  |  |  |
| Non-HDL-C (per 1mg/dL) | 1.0002 (1.000-1.003) | 0.0097 | 1.001 (1.000-1.002) | 0.088 |
| Non-HDL-C (per SD) | 1.075 (1.018-1.135) | 0.0097 | 1.050 (0.99-1.110) | 0.088 |
| *Non-HDL-C, Non-high density lipoprotein-cholesterol; OR, Odds Ratio; CI, confidence interval*<br>**Model 1** adjusted for age, sex, race and education level.<br>**Model 2** adjusted for model 1 plus diabetes, systolic BP, antihypertension medication, body mass index, lipid lowering medications, smoking, and physical activity. |  |  |  |  |

**Supplemental Table 2:** Association of Non-HDL Cholesterol and SCMI Status with CVD Mortality.

| Non-HDL-C and SCMI Status | Participants (n)/ Events (%) | Model 1 |  | Model 2 |  |
| --- | --- | --- | --- | --- | --- |
|  |  | HR (95%CI) | *P-value* | HR (95%CI) | *P-value* |
| SCMI Absent + Low non-HDL-C | 935/8.6 | Reference | -- | Reference | -- |
| SCMI Absent + High non-HDL-C | 4296/11.2 | 0.95 (0.75 – 1.20) | 0.670 | 0.93(0.73 – 1.78) | 0.531 |
| SCMI Present + Low non-HDL-C | 312/16.7 | 1.30 (0.92 – 1.85) | 0.137 | 1.14(0.80 – 1.61) | 0.482 |
| SCMI Present + High non-HDL-C | 1550/19.8 | 1.44 (1.23 – 1.84) | 0.004 | 1.29(1.01 – 1.66) | 0.044 |
| *SCMI = subclinical myocardial injury, non-HDL-C = non-high density lipoprotein-cholesterol, CVD = cardiovascular disease, HR = Hazard Ratio, CI = Confidence Interval*<br>**Model 1** adjusted for age, sex, race and education level.<br>**Model 2** adjusted for model 1 plus history of diabetes, systolic blood pressure, use of antihypertension medication, body mass index, use of lipid lowering medications, smoking status, and physical activity. |  |  |  |  |  |

